# Changes in energy content of menu items at out-of-home food outlets in England after calorie labelling policy implementation: a pre-post analysis (2021-2022)

**DOI:** 10.1101/2024.08.09.24311741

**Authors:** Michael Essman, Thomas Burgoine, Yuru Huang, Andrew Jones, Megan Polden, Eric Robinson, Stephen J. Sharp, Richard Smith, Martin White, Jean Adams

## Abstract

**Importance:** Eating from out-of-home food outlets (OHFO) is common and linked to poor dietary quality, weight gain, and obesity. In response, England implemented mandatory calorie labelling regulations in April 2022 to encourage reformulation and reduce calorie consumption. Few studies have examined the impact of a national calorie labelling policy on OHFO menus.

**Objective:** Examine pre-post changes in energy content of menu items from large OHFO in England after mandatory calorie labelling.

**Design, Setting, and Participants:** Observational study using data from MenuTracker, a longitudinal database of online menus. Data were collected in September 2021 (pre-policy) and September 2022 (post-policy) from large OHFO in England. 15,057 pre-policy and 15,988 post-policy menu items were included from 78 large OHFO chains present in both periods.

**Intervention:** Implementation of mandatory calorie labelling on menus of large OHFOs in England.

**Main Outcomes and Measures:** Mean energy content (kcal) of menu items, examined overall and by food group and chain type. Changes in energy content for removed, continuous, and new items to assess reformulation.

**Results:** Overall, a reduction of 9 kcal (95% CI: −16 to −1) in mean energy content was observed post-policy. Significant reductions per item in beverages (−36 kcal), burgers (−103 kcal), and mains (−30 kcal). By chain type, significant reductions per item in pubs, bars, and inns (−52 kcal), restaurants (−23 kcal), and entertainment venues (−49 kcal). Changes driven by removal of higher kcal items (458 kcal, 95% CI: 394 to 523) and addition of lower kcal items (434 kcal, 95% CI: 370 to 499). No significant change in energy content for continuously available items, indicating limited evidence of reformulation.

**Conclusions and Relevance:** The 2022 mandatory calorie labelling policy in England led to a small reduction in mean energy content of menu items, driven by removal of higher calorie items and addition of lower calorie items. Elsewhere, we did not find evidence of changes in kcal purchased or consumed, suggesting these menu changes did not focus on the most commonly consumed items. Further research is needed to evaluate longer-term menu changes and additional strategies to enhance policy impact on consumer behavior and public health.

## Introduction

Eating food from out-of-home food outlets (OHFO) is common and associated with poorer dietary quality, weight gain and obesity, posing significant challenges to public health (1–3). A critical issue in unhealthy out-of-home eating is the tendency of individuals to underestimate their energy consumption, particularly when exposed to the high energy-density foods commonly offered by OHFO (1,4,5). A study using 2018 data from UK chains with 50 or more locations found 96% of meals in full-service restaurants and 70% in fast-food outlets exceeded 600 kcal (6). Recognizing the urgency of addressing the public health implications of out-of-home food consumption, the UK Government implemented mandatory calorie labelling regulations on April 6th, 2022 (7). These regulations require large OHFOs in England, defined as those with 250 or more employees, to display kilocalorie (kcal) information for menu items at the point of sale. The regulations apply to food and non-alcoholic drink items ready for immediate consumption, excluding pre-packaged foods and specifically exempted items, such as unprepared fruits and vegetables, single ingredient unprocessed foods, and others (8).

Despite the potential of such labels to reduce energy purchased, the evidence for the effect of energy labels on customer selections is mixed. Studies conducted in the United States present varying results, with some suggesting a reduction in daily caloric intake, while others find no significant effect on energy content ordered or consumed (9–11). In England, we found no evidence of changes in kcal purchased or consumed post-implementation (12). Furthermore, the type of food outlet may influence customer behaviour, with greater reductions observed in cafes and sit-down restaurants compared to fast-food outlets (13). These results could be explained by different customer expectations for different out-of-home (OOH) eating occasions. Qualitative work in the OOH food sector in England suggests customer expectations could be a key barrier to the impact of menu labelling on ordering lower energy items (14).

Energy labelling policies may not only act as an information intervention to change customer behaviour, but may also incentivise retailers to change the energy content of menu items if businesses do not want to report selling excessively high calorie items (14). Recent qualitative work suggests that large OHFOs may take a “health by stealth” approach to reformulation: gradually reducing the energy, sugar, and salt content of their products as a response to labelling regulations to make food options slightly healthier without being noticed or disapproved of by customers (14). Menu changes can take the form of reformulating existing products, discontinuing higher energy products, or adding lower energy products. All three methods could reduce the mean kcal content of OHFO menus. A recent meta-analysis of 45 studies found that mandatory energy labelling was associated with a 15 kcal reduction in menu items offered (11). These reductions could potentially translate into lower energy intake by customers if they select lower-energy products or consume reformulated items.

Before mandatory labelling policies were implemented in the United States and England, there was some evidence that restaurants that implemented voluntary energy labelling sold lower fat and salt items than those without such labelling (15,16). However, this finding could reflect a preference for labelling amongst those selling healthier items. There is less evidence that demonstrates that mandatory labelling reduces energy content. Most of the previous evidence for national-level energy labelling policies is from the United States (17–20), where a recent study found that mandated energy labelling may have encouraged large restaurant chains to introduce lower-energy items (21). Another study of locally implemented labelling policies in the USA found no changes in mean energy on menus between experimental and control fast-food chains but did find healthier items at the labelled locations (22). Although meta-analyses suggest small but potentially beneficial improvements to menus after energy labelling (23), customers may not benefit from average reductions if they do not select lower energy products. Therefore, it is essential to identify which food categories are most subject to change to determine where dietary improvements can be made. Different types of food businesses sell different food items that may vary in the degree to which they can be feasibly reformulated. Therefore, examining any differences by type of food business may also be informative.

To address these research gaps, this study examines pre-post changes in the energy content of menu items before and after the calorie labelling (Out of Home Sector) (England) regulations. We also examine changes in energy content across different food categories and types of food businesses, offering insights into where reformulation and menu changes most occurred.

## Methods

Using the MenuTracker database, the first longitudinal nutritional database of food prepared out of the home in the UK (24), we assessed both reformulation and menu changes from before to after introduction of the labelling regulations. We defined reformulation as changes in the same *continuously available* item at both time periods. Menu changes were defined as removals of items from the pre-period or addition of new items to the post-period. We examined changes in mean kcal content of new, removed, and continuously available items, as well as changes in mean kcal by food category and chain type. Additionally, we examined the proportions of menu items exceeding recommended energy intake per meal (>600 kcal) according to current guidance in England (25). These analyses were conducted for chains that were present in the database at both time periods ie ‘core chains’. Finally, we conducted a full landscape analysis using all available data from all available chains at both time periods.

### Data source

Data in the longitudinal database MenuTracker were collected using web scraping techniques and PDF extraction tools from food businesses that posted calorie information for menu items online and were subject to the regulations (24). If available on business websites, MenuTracker collects food item name and description, serving size, energy, macronutrients, fibre, salt, allergens, special dietary information, and menu section (used to determine whether items are children’s items or sharing items). We used two waves of data, collected 12 months apart in September 2021 (pre-policy) and September 2022 (post policy) to minimise any effects of seasonal variation. A single scrape of data collection was done for each chain website and ran from August 25 to 31, 2021 for the September 2021 data collection and from August 17 to September 3 for September 2022 data collection. Using data collected in September 2021 to represent the pre-policy period also allowed us to minimise the effect of early changes associated with implementation in April 2022 that might have occurred during the six months before the policy came into force.

MenuTracker September 2021 (pre-period) data included 79 unique out-of-home food businesses (henceforth referred to as chains) subject to the regulations. MenuTracker September 2022 (post-period) included 90 chains. One chain from the pre-period did not present information online in the post-period, and therefore our main analysis includes the 78 chains operating at both time periods. For simplicity, these 78 chains will be referred to as *core chains* (Supplementary Table 1). We also conducted a *full landscape* analysis using all available data from all chains at both time periods (i.e. 79 pre and 90 post). This expanded database is described in Supplementary Table 2.

### Sample Size

The pre-period data included 19,392 menu items. After removing 1,937 duplicate entries, the dataset was reduced to 17,455 menu items. Where items had missing kcal information but had full macronutrient information (values for protein, carbohydrates and fats), these macronutrient values were used to generate kcal values based on 4 kcal per gram of protein, 4 kcal per gram of carbohydrates, and 9 kcal per gram of fat. After this replacement for 59 items, 2,370 items were deleted due to having no kcal information, resulting in 15,085 unique items with kcal information. The post-period data initially included a total of 24,097 menu items. After removing 4,795 duplicate entries and deleting 1,406 items with no kcal information, there were 17,896 unique items with kcal information remaining. Thus, our full landscape analysis consisted of a total of 32,981 items (Figure 1). When restricting the analysis to only the 78 chains present at both time periods, there were 15,057 observations in the pre-period and 15,988 observations in the post-period, totalling 31,045 items (Figure 2).

**Figure 1.**
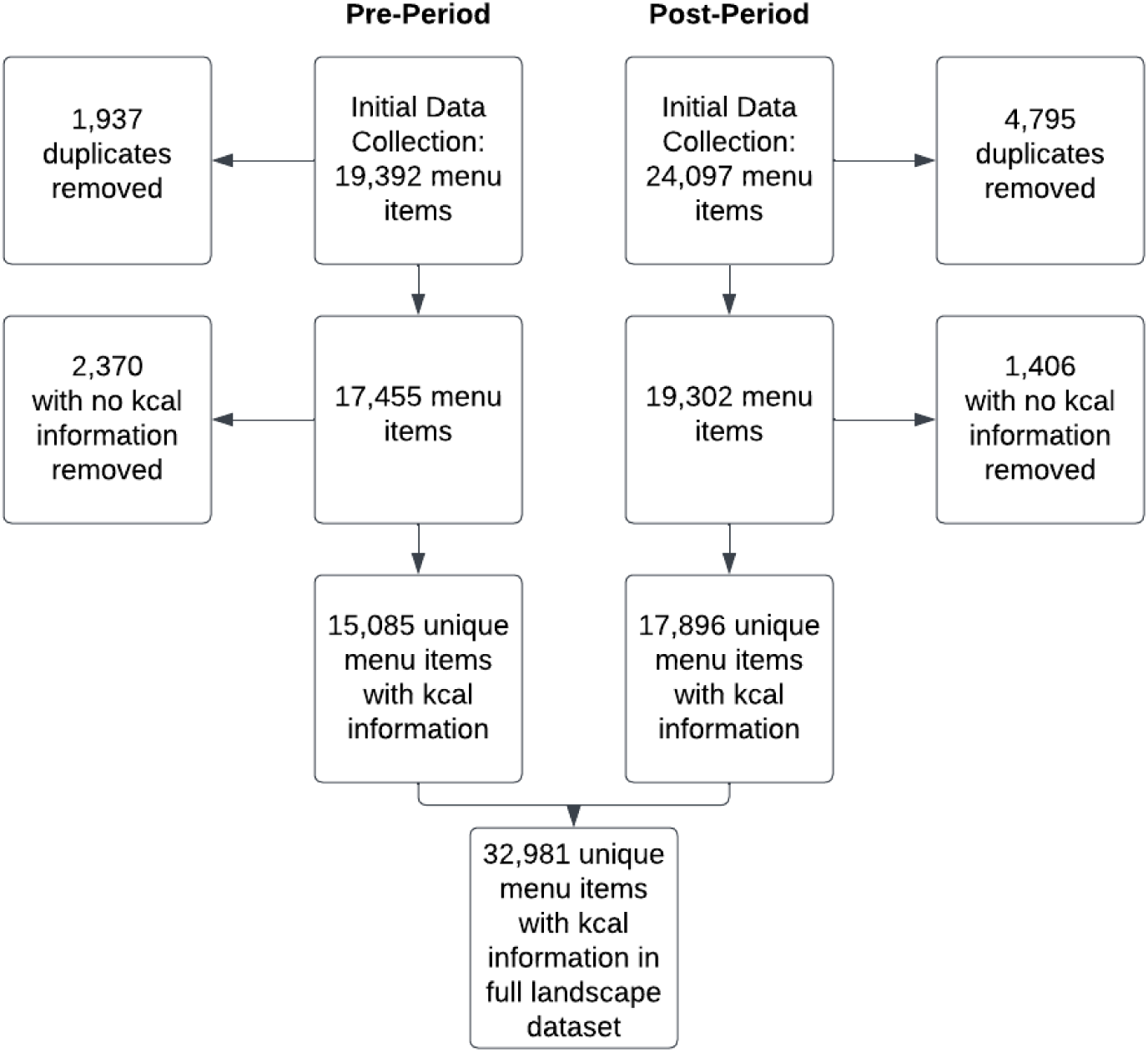
Flowchart for data collection for full landscape analysis: all 90 available chains included.

**Figure 2.**
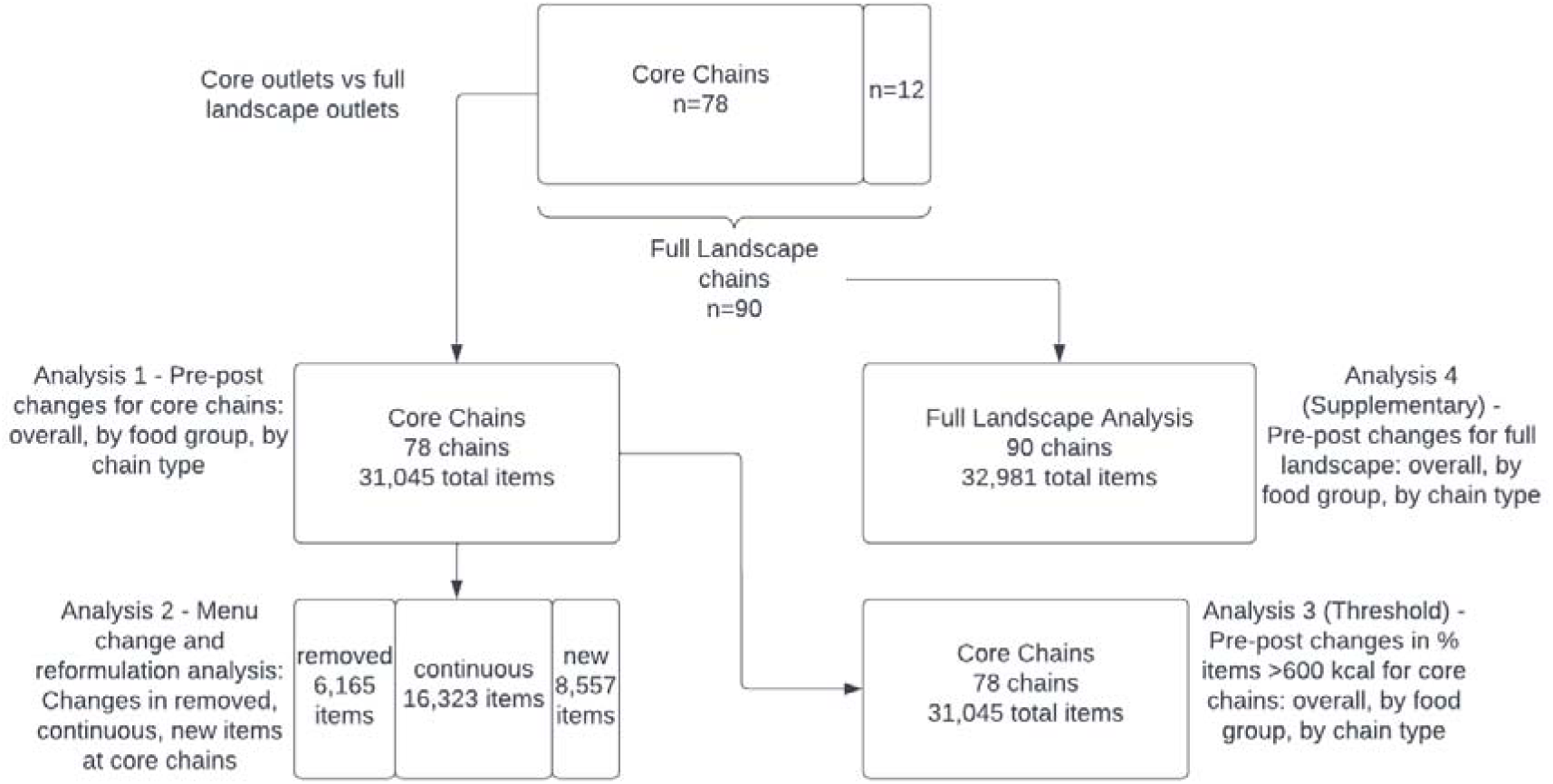
Conceptual diagram for analyses, including total number of chains and items

### Food Groups

Menu items were classified via a semi-automated process by ME into 12 food groups used in previous work in both the United States and United Kingdom (15,26,27) (Table 1). Following previous work, all variations on items were included as separate items (e.g. cappuccino made with full fat cow’s milk, semi-skimmed cow’s milk, coconut milk, and soy milk count as four separate items) (15). A reliability check of food group assignments was conducted on a 2% random sample of items (n=648) with 91% agreement and coding decisions resolved between the two coders (ME and YH).

**Table 1.** Food groups used in analysis, with descriptions. Adapted from previous work (15).

| **Food group** | **Description and examples** |
| --- | --- |
| Appetisers and sides | Items that are listed in “appetisers,” “starters,” or “sides” menu sections. Also includes items or small dishes served to complement the main course, for example a chicken skewer “add on” to a burger or meal, or a “side of vegetables/rice/beans/fruit/etc.” |
| Baked goods | Food items prepared by baking. Examples include bread, cakes, cookies, and pastries. |
| Beverages | Menu items intended for drinking, including both non-alcoholic and alcoholic options. Examples include water, tea, coffee, juice, and soft drinks. |
| Burgers | All items described on menus as burgers. Examples include burger, hamburger, chicken burger, veggie burger. |
| Desserts | All sweets intended to be served as desserts. Also includes baked goods served as desserts (e.g. cakes) and candy. Other examples include cookies and ice cream. |
| Fried potatoes | Potatoes that have been cooked by frying, often in oil. Examples include French fries, sweet potato fries, potato chips, and hash browns. |
| Mains | The primary or most substantial item in a meal, usually featuring a significant source of protein. Examples include steak, chicken curry, grilled fish, chicken nuggets. |
| Pizza | A dish consisting of a flatbread base typically topped with sauce, cheese, and various toppings. Includes items listed as pizzas or flatbreads. |
| Salads | Dishes composed primarily of raw or cooked vegetables, often mixed with other ingredients and served with a dressing. Examples include garden salads, side salads, or Caesar salads. |
| Sandwiches | Foods consisting of one or more types of filling, such as meat, cheese, or vegetables, placed between slices of bread or rolls. Examples include ham sandwiches, subs, and wraps. |
| Soups | Liquid dishes typically made by simmering ingredients like meat, vegetables, and legumes in broth or water. Examples include chicken noodle soup, tomato soup, and minestrone. |
| Toppings and ingredients | Items added to enhance the flavour of a dish, including condiments, garnishes, and additional components. Examples include cheese, croutons, sauces, dressings. Also includes beverage toppings such as whipped cream added to coffee. |

### Chain types

Chains were classified into six chain types: cafes and bakeries (henceforth referred to as cafes); Western fast food and takeaways; pubs, bars, and inns; restaurants; sports and entertainment; and Asian Fast Food based on how chains described their outlets and food offerings on their websites. All chains are listed within each category in Supplementary Tables 1 and 2.

### Removed, continuous, and new items

To distinguish reformulation from other menu changes, we categorized all items at the 78 *core chains* into *removed*, *continuous*, and *new* items. *Continuous* items were defined as items with the same name at the same outlet present in both pre- and post-periods. We defined *removed* items as present in the pre-period but not the post-period, and we defined *new* items as present in the post-period but not pre-period.

Probabilistic record linkages were conducted according to published best practices, followed by manual checking, to identify items with the same name at the same period at both time points (28). Matches required the identical chain name, but item names could be a fuzzy (ie similar but imperfect) match.

We used the ‘reclink’ package in Stata to facilitate probabilistic record linkage. The ‘reclink’ command uses matching algorithms to determine the likelihood of a match between records based on the similarity of item names (29). This algorithmic approach was followed with manual checking by ME. There were two stages of manual checks. The first step was to check for false positive matches using the clrevmatch reviewing tool (28), which juxtaposes all potential matches next to each other and allows the user to confirm whether it is a correct match. An example of a potential false positive that can occur during probabilistic matching under conditions of high similarity would be an erroneous match between “Caramel Macchiato Coconut Milk 12 oz” and “Caramel Macchiato Coconut Milk 16 oz” despite only a single character difference. Thus, highly similar, but not exact, matches were manually checked to ensure they were true matches. The second manual check was to identify false negative matches that were missed during probabilistic matching. ME manually checked the entire database sorted first by chain name and then by item name to identify potential matches that were not captured by the fuzzy match. These cases occurred if the outlet changed the way the item name was recorded between pre- and post-periods, even if it was clearly the same item. For example, despite less character similarity than the macchiato example, “Beetroot Latte with coconut” is a match with “Beetroot Latte coconut milk”.

A further manual check was conducted to explore reliability of the new, continuous, and removed designations. A random chain was selected from each chain type to account for possible menu differences, resulting in 2,471 items from a total of 32,981 (7.5%). There was high agreement between the coders (96.2%).

## Analysis

All statistical analyses were conducted in Stata 17. We conducted four analyses, displayed in the concept map (Figure 2) for clarity and described in greater detail below. These analyses were:

1. Estimate pre-post differences in mean energy (kcal) at *core chains* (n=78) overall, by food group, and by chain type.
2. Estimate pre-post differences in mean energy (kcal) for each of removed, continuous, and new items at *core chains* (n=78) overall, by food group, and by chain type.
3. Estimate pre-post differences in prevalence of items that exceed 600 kcal threshold at *core chains* (n=78) overall, by food group, and by chain type.
4. Estimate pre-post differences in mean energy (kcal) using the *full landscape* of chains (n=90) overall, by food group, and by chain type.

We first detail the general modelling approach for Analysis 1 including covariates, followed by adaptations to this for Analyses 2-4.

### Analysis 1: Estimate pre-post differences in mean energy (kcal) at core chains (n=78) overall, by food group, and by chain type

We used linear mixed regression models with random intercepts to estimate the mean energy content (kcal) for items overall, by food group, and by chain type. Models included a three-level hierarchical structure to account for inherent clustering of data: menu items represented the first level (level 1), nested within chains (level 2), which were further nested within chain types (level 3). Models were adjusted at the item-level for children’s menu item status, sharing items and food group. Including item-level covariates adjusts for differences in the types of food items sold across outlets because sharer platters, children’s menus, and food groups differ in calorie content and size and are therefore associated with energy content. We also conducted a sensitivity analysis that removed food group from the adjustment set, which may more closely reflect what customers see on menus. We estimated mean kcal and 95% confidence intervals (CIs) for the pre- and post-policy periods for items overall using average marginal effects (AME), and two-way interactions between time (binary) with food group and with chain type to estimate mean kcal and 95% CIs for each combination of time and category. Pairwise comparisons of margins were conducted to assess differences between time periods for each level of the categorical variable, and each contrast is presented with 95% CIs.

### Analysis 2: Estimate pre-post differences in mean energy (kcal) for each of removed, continuous, and new items at core chains (n=78) overall, by food group, and by chain type

For the second analysis, we used the same modelling approach as outlined above, but also included an indicator variable for whether items were new, continuous or removed. We estimated the marginal mean kcal and 95% confidence intervals (CIs) for removed items, new items, and for continuous items. Pairwise comparisons of margins were conducted to assess differences for removed versus continuous, new versus continuous, and new versus removed. We interpreted the pre-post changes in continuous items as an analysis of reformulation.

### Analysis 3 (Threshold): Estimate pre-post differences in prevalence of items that exceed 600 kcal threshold at core chains (n=78) overall, by food group, by chain type, and by new, continuous, and removed

Our third analysis examined the proportion of items that exceeded England’s per meal recommendations (>600 kcal) before and after the policy using the same three-level hierarchical structure but in this case mixed effects logistic regression models for the binary outcome (25). We estimated the marginal probability of this outcome (>600 kcal) by overall menu items, food group, and chain type at each time period.

### Analysis 4 (Supplementary): Estimate pre-post differences in mean energy (kcal) using the full landscape of chains (n=90) overall, by food group, and by chain type

The final analysis was a full landscape analysis using all available data, which includes all items from chains that are included in MenuTracker and posted calorie information online either pre- or post-policy. Total chains included 79 pre-policy and 90 post-policy. We followed the same modelling approach as the first analysis, with a three-level hierarchical structure and the same item level covariate adjustments. We also estimated the same two-way interaction marginal effects for each category of food type over time and each category of chain type over time.

## Results

### Descriptive statistics for item availability at each data collection wave

Descriptions of item availability in each data collection wave (pre-policy Sept 2021 and post-policy Sept 2022) at core chains and full landscape chains are presented in Table 1. There were approximately two thousand more menu items present in the full landscape analysis that the core chain analysis (Table 1). The most common food groups at both pre- and post-policy were beverages and mains, and the least common items were soup and fried potatoes (Table 1). The number of items at each chain as well as the chain type classifications are presented in Supplementary Tables 1 and 2.

**Table 1.** Summary statistics for item availability at core chains (n=78) and full landscape chains (n=90)

|  | Core Chains |  | Full Landscape |  |
| --- | --- | --- | --- | --- |
| □ | Pre-Policy<br>(Sept 2021) | Post-Policy<br>(Sept 2022) | Pre-Policy<br>(Sept 2021) | Post-Policy<br>(Sept 2022) |
| Chains, n | 78 | 78 | 79 | 90 |
| Menu items, n | 15,057 | 15,988 | 15,085 | 17,896 |
| **Menu items by food group, n(%)** |  |  |  |  |
| Appetisers and sides | 1869 (12.4) | 2409 (15.1) | 1869 (12.4) | 2824 (15.8) |
| Baked goods | 397 (2.6) | 451 (2.8) | 410 (2.7) | 519 (2.9) |
| Beverages | 3474 (23.1) | 4110 (25.7) | 3474 (23.0) | 4454 (24.9) |
| Burgers | 411 (2.7) | 388 (2.4) | 411 (2.7) | 490 (2.7) |
| Desserts | 1164 (7.7) | 1493 (9.3) | 1171 (7.8) | 1642 (9.2) |
| Fried potatoes | 222 (1.5) | 202 (1.3) | 222 (1.5) | 228 (1.3) |
| Mains | 3427 (22.8) | 2926 (18.3) | 3428 (22.7) | 3461 (19.3) |
| Pizza | 2093 (13.9) | 2008 (12.6) | 2086 (13.8) | 2044 (11.4) |
| Salads | 338 (2.2) | 439 (2.8) | 338 (2.2) | 494 (2.8) |
| Sandwiches | 796 (5.3) | 905 (5.7) | 801 (5.3) | 1015 (5.7) |
| Soup | 102 (0.7) | 110 (0.7) | 102 (0.7) | 133 (0.7) |
| Toppings and ingredients | 764 (5.1) | 547 (3.4) | 771 (5.1) | 587 (3.3) |
| **Menu items by chain type, n (%)** |  |  |  |  |
| Cafes | 3792 (25.2) | 3558 (22.3) | 3792 (25.1) | 3861 (21.6) |
| Fast food and takeaway | 4136 (27.5) | 3813 (23.9) | 4164 (27.6) | 3879 (21.7) |
| Pubs, bars, and inns | 3584 (23.8) | 4071 (25.5) | 3584 (23.8) | 4735 (26.5) |
| Restaurants | 2412 (16.0) | 3045 (19.1) | 2412 (16.0) | 3830 (21.4) |
| Sport and Entertainment | 901 (6.0) | 1164 (7.3) | 901 (6.0) | 1254 (7.0) |
| Asian Fast Food | 232 (1.5) | 337 (2.1) | 232 (1.5) | 337 (1.9) |

### Analysis 1: Estimate pre-post differences in mean energy (kcal) at core chains (n=78) overall, by food group, and by chain type

Figure 3 presents mean kcal content for items at core chains, both before and after the implementation of the policy, overall, by food group, and by chain type, and Figure 4 presents pre-post differences for items at core chains overall, by food group, and by chain type. Prior to the policy, the estimated mean kcal for all items was 445 kcal, and following the policy, the estimated mean kcal decreased to 436 kcal, a 9 kcal (95% CI: −16 to −1) reduction (Fig. 3). According to the sensitivity analysis that removed food group from the adjustment set, which may more closely reflect what customers see on menus, overall mean kcal was 446 (95% CI 357 to 535) in the pre-policy period and 424 (95% CI 336 to 513) in the post-policy period, a difference of 21 kcal (95% CI −30 to −12).

**Figure 3.**
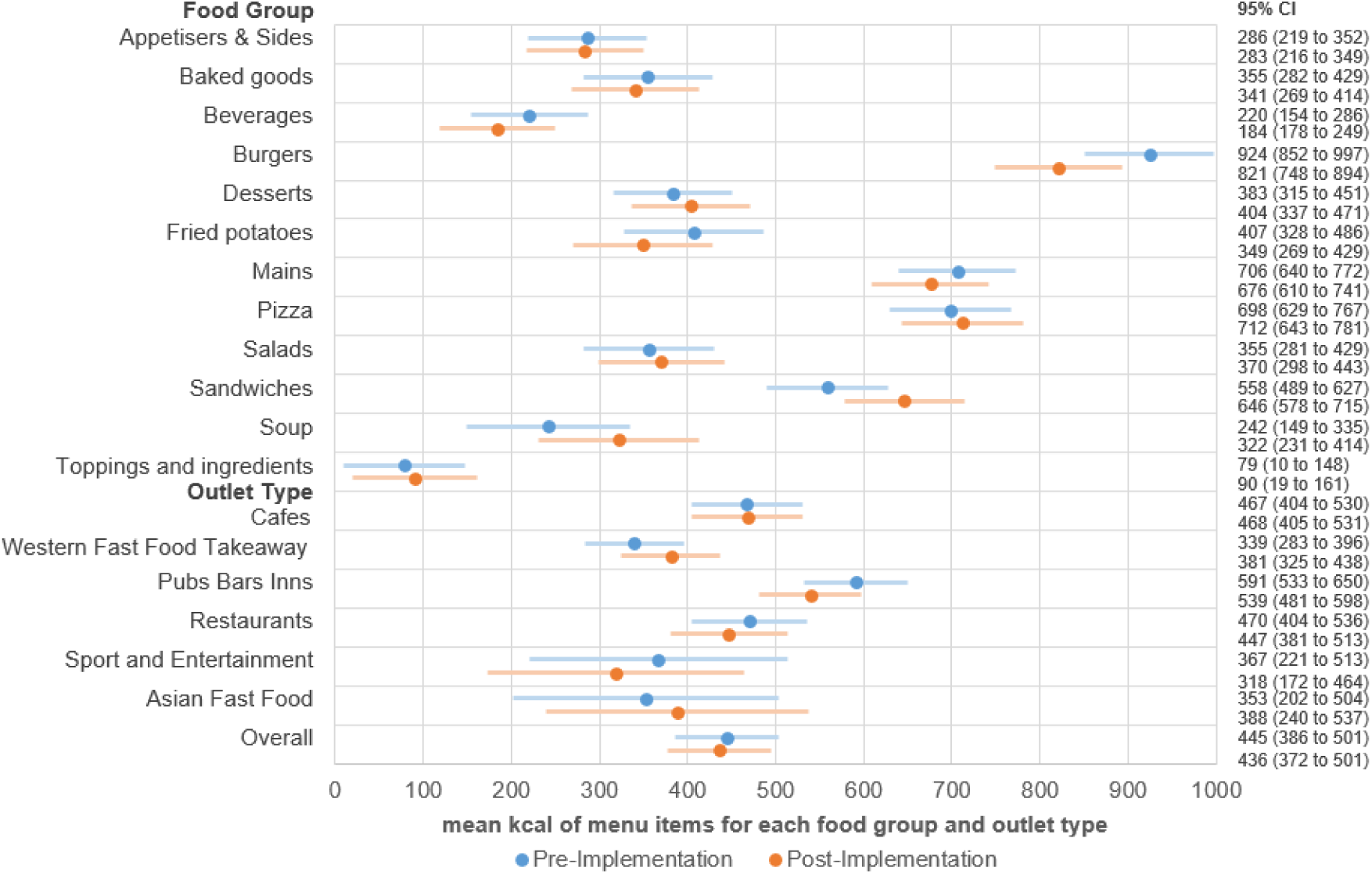
Marginal mean kcal from linear mixed model overall, by food group, and by chain type for all items available at core chains (n=78) using MenuTracker data from pre- (September 2021) and post-policy (September 2022), total n items=31,045

**Figure 4.**
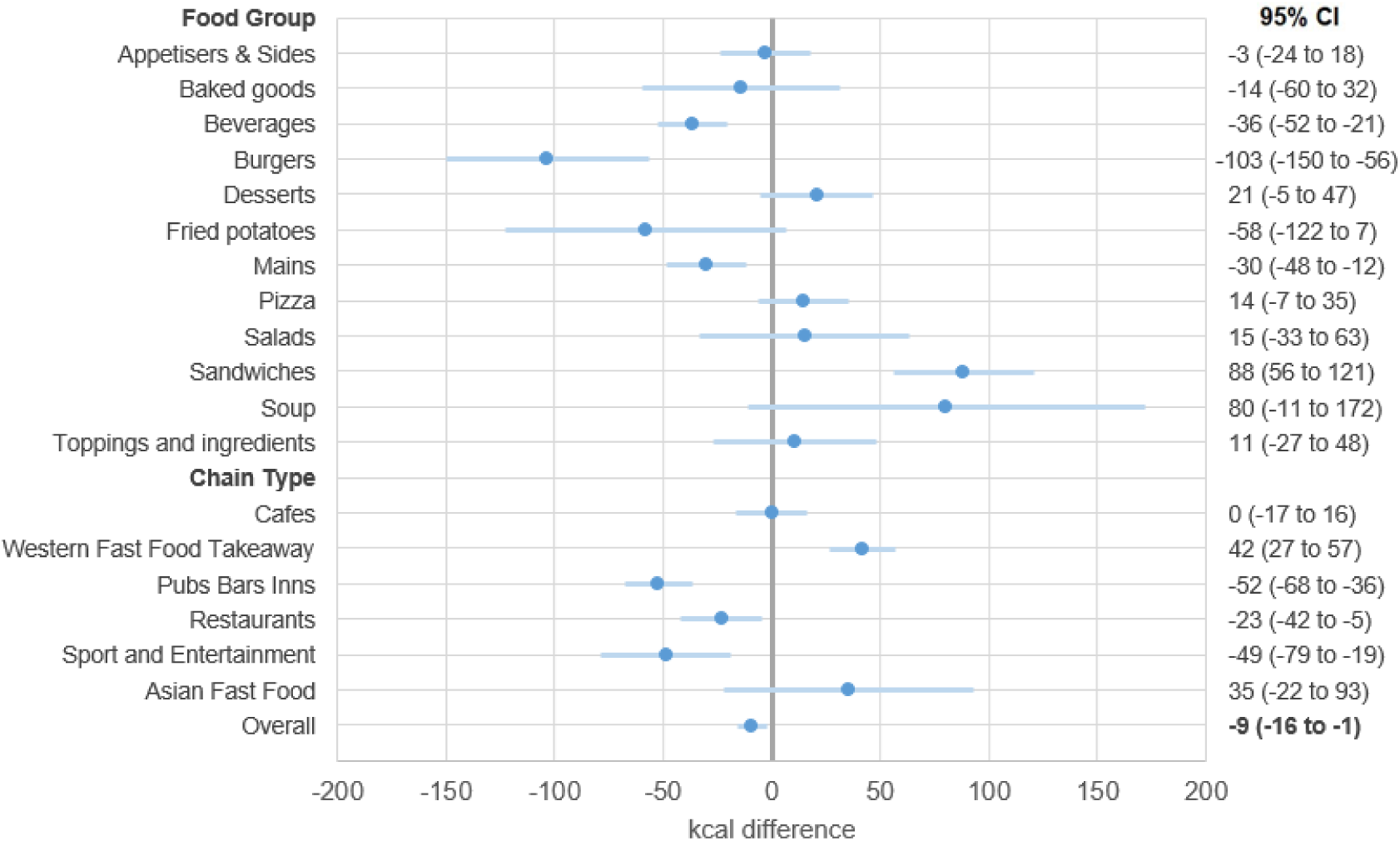
Pre-post differences in kcal overall, by food group, and by chain type estimated from linear mixed model for all items available at core chains (n=78) using MenuTracker data from pre- (September 2021) and post-policy (September 2022), total n items=31,045

The highest kcal per item food groups were burgers, mains, and pizzas, and the highest kcal per item chain types were restaurants, and pubs, bars, and inns (Fig. 3). After the policy, the largest significant reductions were 103 fewer kcal (95% CI: −150 to −56) for burgers, 36 fewer kcal (95% CI: −52 to −21) for beverages, and 30 fewer kcal (95% CI: −48 to −12) for mains (Fig. 4). There were few soups in either time period, leading to a large confidence interval in the change (Fig. 4). Sandwiches increased by 88 kcal (95% CI: 56 to 121) pre-post (Fig. 4). When analysed by chain type, statistically significant results included a reduction of 52 kcal (95% CI: −68 to −36) at pubs, bars, and inns, a reduction of 38 kcal (95% CI: −42 to −5) for restaurant items, and a reduction of 49 kcal (95% CI: −79 to −19) for Sports and Entertainment items. There was an increase of 42 kcal (95% CI: 27 to 57) for Western Fast Food and Takeaway items (Fig. 4).

### Analysis 2: Estimate pre-post differences in mean energy (kcal) for removed, continuous, and new items at core chains (n=78) overall, by food group, and by chain type

Figure 5 presents estimated mean kcal content from the linear mixed model for items categorized as removed, new, and continuously available, before and after the policy implementation. Prior to the policy, removed items had an estimated mean energy content of 458 (95% CI: 394 to 523) kcal, and continuously available items had 437 (95% CI: 373 to 501) kcal. After the policy, new items had 434 (95% CI: 370 to 499) kcal, and continuously available items had 439 (95% CI: 374 to 503) kcal, no change compared to pre-policy (Fig. 5). Removed items contained 21 (95% CI: 8 to 34) more kcal than continuous items and 25 (95% CI: 9 to 41) more kcal than new items (Fig. 6).

**Figure 5.**
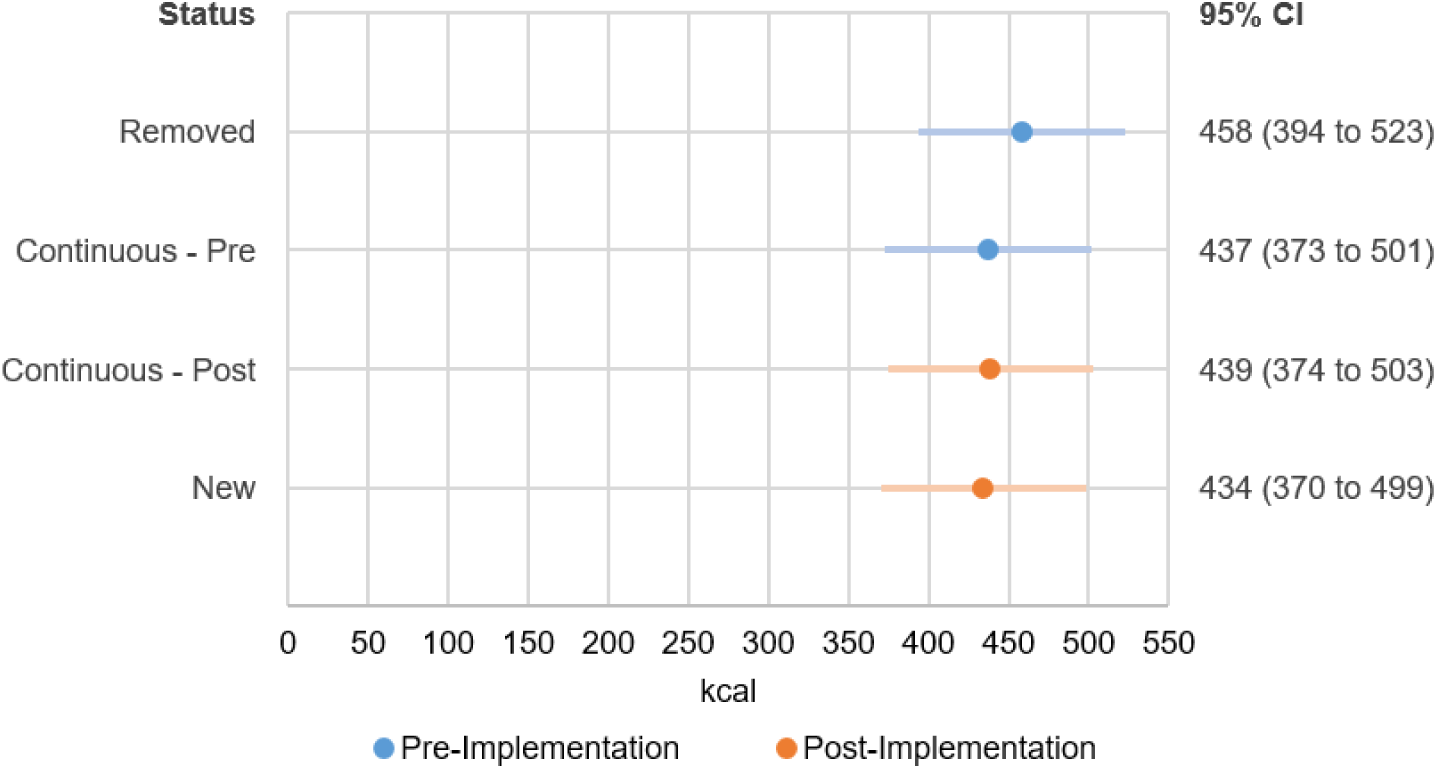
Marginal mean kcal for removed, continuous, and new items estimated from linear mixed model at core chains (n=78) using MenuTracker data from pre- (September 2021) and post-policy (September 2022), total n items=31,045

**Figure 6.**
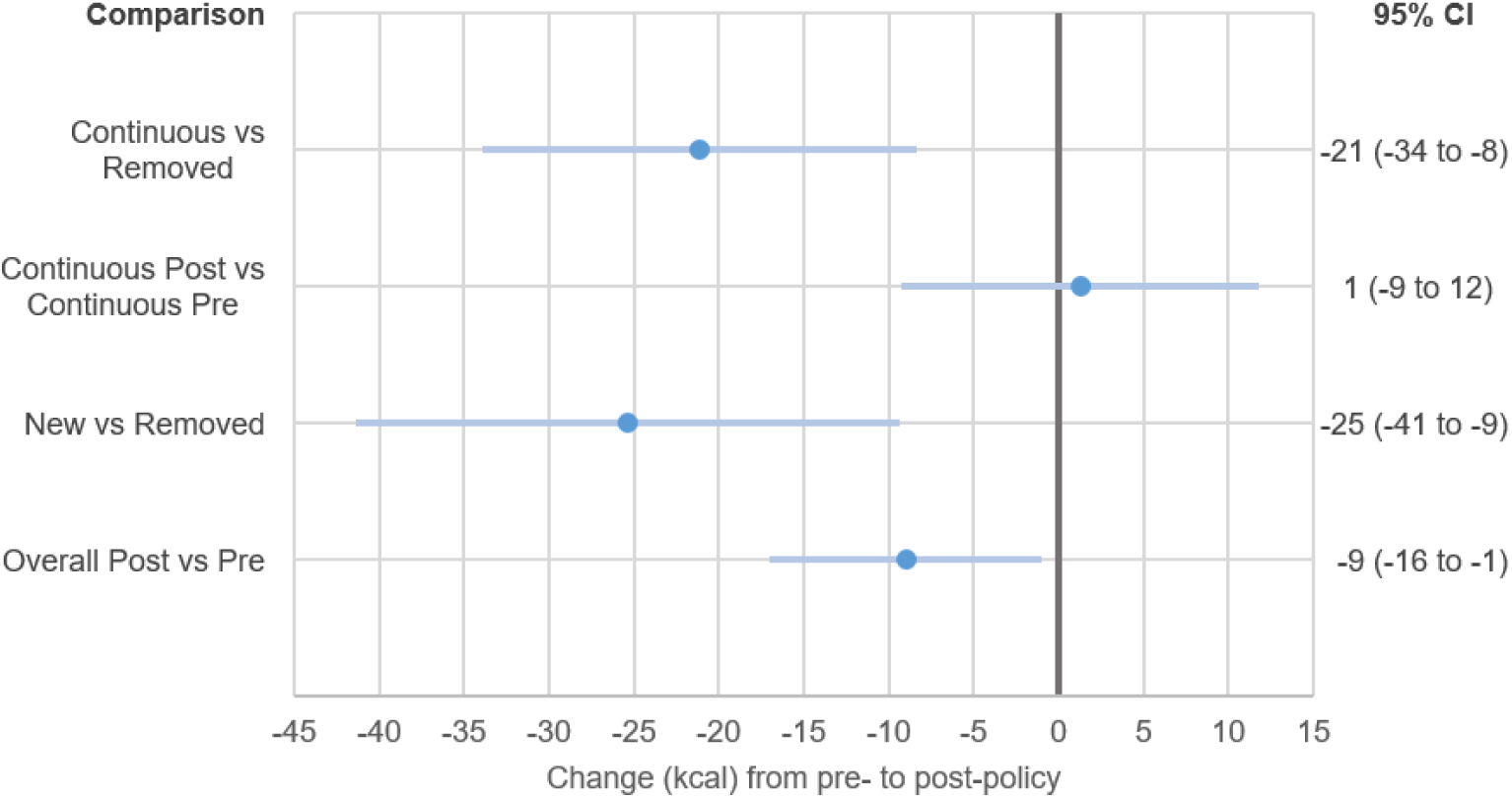
Differences in kcal for comparisons of removed, new, and continuous items estimated from linear mixed model at core chains (n=78) using MenuTracker data from pre- (September 2021) and post-policy (September 2022), total n items=31,045

### Analysis 3 (Threshold): Estimate pre-post differences in prevalence of items that exceed 600 kcal threshold at core chains (n=78) overall, by food group, by chain type, and by new, continuous, and removed

Figure 7 presents the proportion of items over the 600-kcal threshold for core chains, both before and after the implementation of the policy. Prior to the policy, 21.8% of items (95% CI: 15.3% to 28.3%) were over 600 kcal, and after the policy, 22.2% of items (95% CI: 15.6% to 28.7%) were over 600 kcal, with no difference between pre- and post-policy.

**Figure 7.**
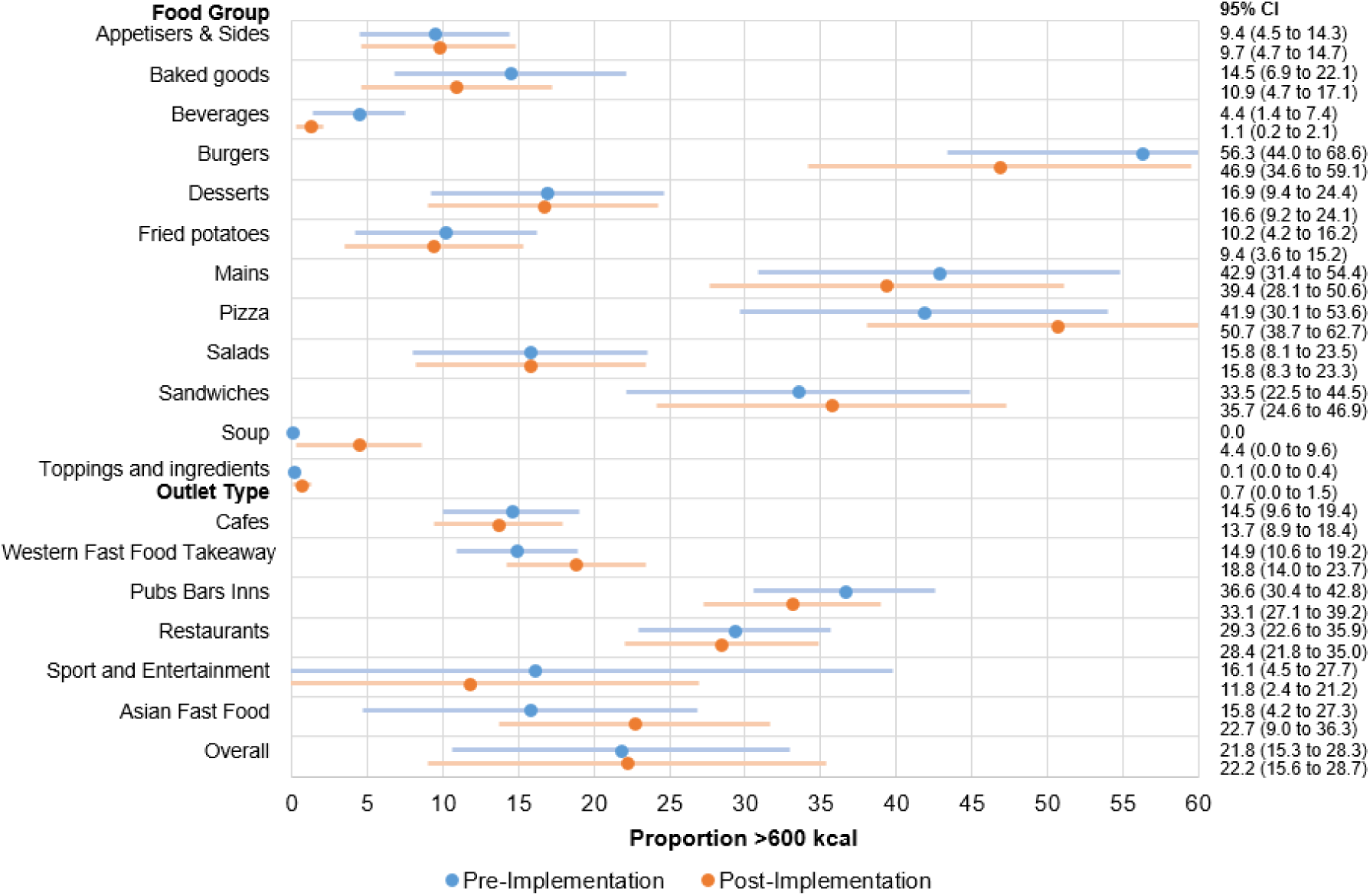
Proportion of items that exceed 600 kcal estimated from linear mixed model overall, by food group, and by restaurant type for all items available at core chains (n=78) using MenuTracker data from pre- (September 2021) and post-policy (September 2022), total n items=31,045

The pattern of results for analysis 3 largely reflected analysis 1. The food groups with the most items over 600 kcal were burgers, mains, and pizzas, and the chain types with the most items over 600 kcal were restaurants and pubs, bars, and inns (Figure 7). After the policy, the proportion of items exceeding 600 kcal was 3.2% (95% CI: −5.6 to −0.8) lower for beverages, 9.5% (95% CI: −15.6% to −3.4%) lower for burgers, and 3.4% (95% CI: −5.6% to - 1.3%) lower for mains. The proportion of items exceeding 600 kcal increased by 9.0% (95% CI 6.4% to 11.6%) for pizzas (Figure 8). When analysed by chain type, the proportion of items exceeding 600 kcal was 3.9% (95% CI: −5.7% to −2.2%) lower at pubs, bars, and inns (Figure 8).

**Figure 8.**
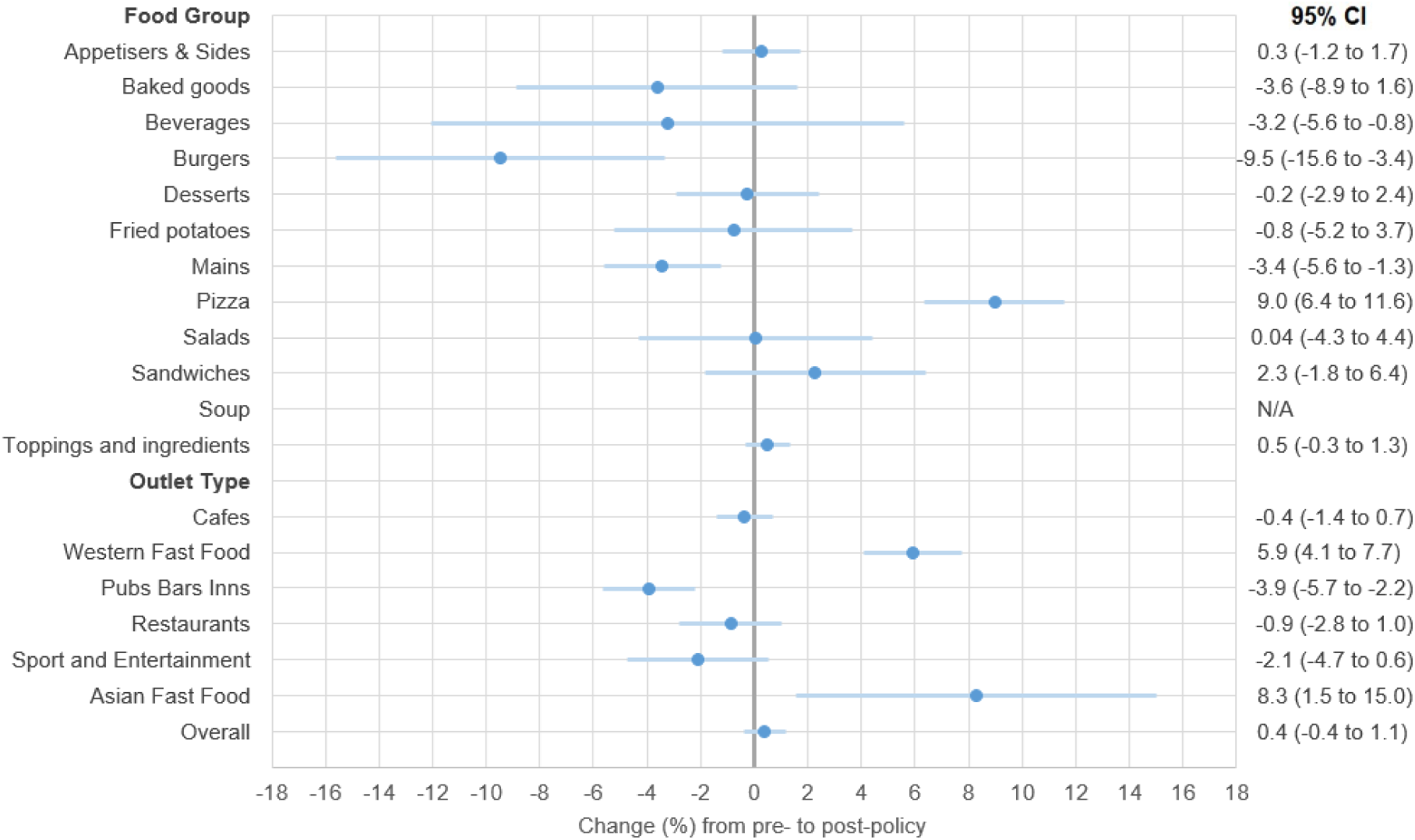
Pre-post differences in proportion of items that exceed 600 kcal estimated from linear mixed model overall, by food group, and by chain type for all items available at core chains (n=78) using MenuTracker data from pre- (September 2021) and post-policy (September 2022), total n items=31,045

### Analysis 4: Estimate pre-post differences in mean energy (kcal) using the full landscape of chains (n=90) overall, by food group, and by chain type

The results from the full landscape analysis are presented in Supplementary Table 3. The overall mean kcal was 458 (95% CI 412 to 505) in the pre-policy period and 450 (404 to 497) in the post-policy period, and difference of 8 fewer kcal (−8 to 0.2) in the post-period that was not statistically significant. The overall pattern of results reflected Analysis 1, with lower kcal values in beverages, burgers, and mains, lower kcal values at restaurants and pubs, bars, and inns, and an increase in kcal offered by Western fast-food chains.

## Discussion

### Statement of principal findings

To our knowledge, this study is the first to examine menu changes at out of home food chains in relation to implementation of the 2022 national calorie labelling policy. Our study examining changes in online menu information from large out-of-home food chains in England found a small (−9 kcal, 95% CI: −16 to −1) reduction in mean kcal after implementation of the calorie labelling policy compared to before. Models not adjusted for food group changes, which indicate what customers may perceive on menus without accounting for food group distributions, showed a greater difference than adjusted findings, with a −21 kcal lower (95% CI −30 to −12) mean after policy implementation. These mean changes were driven by the removal of higher calorie items on menus. There were no pre-post changes in kcal for continuously available items. Thus, we find some evidence for changes in menus but limited evidence for reformulation of existing menu items. The greatest changes in food groups were for burgers, beverages, and mains. When analysed by chain type, the significant changes included reductions in kcal of items at pubs, bars, and inns, restaurants; and sports and entertainment chains and an increase at Western fast food and takeaway chains. The food groups with the most items over 600 kcal were burgers and pizzas, and the chain types with the most items over 600 kcal were restaurants and pubs, bars, and inns. The full landscape analysis found a similar pattern of results to the core chain analysis.

### Interpretation of findings

The small reduction in average kcal of items available on menus we found is likely to have modest to limited impact on population health. Indeed, another pre-post study of the labelling regulations using approximately 3,000 customer intercept surveys found no change in kcal purchased or consumed (12). It is also possible that chains already reformulated some menu items between the announcement of the calorie labelling policy, which was approved in July 2021, and the initial data collection for this study, in September 2021.

We identified some evidence of differential pre-post changes by food group and chain type. In general, food groups with higher baseline kcal such as burgers and mains had greater absolute reductions. One possible explanation for this is that changing larger kcal items is easier due to a larger baseline portion size or high calorie density. Given the high energy density of fat, reducing the fat content of high-calorie, particularly high-fat items, could substantially reduce the energy density of the food (30). Another explanation for the reductions in larger food groups could be that there is some potential embarrassment in having extremely high kcal items on menus once labelling is mandatory (14). It may be easier to reduce mean kcal offered in the beverage category due to the availability of low and zero calorie options, which are typically not feasible for foods. The larger absolute changes apparently associated with larger baseline kcal levels also applies to chain type, with significant reductions observed for restaurants as well as pubs, bars, and inns and for sport and entertainment venues, which mostly comprised of cinemas.

We found more evidence of menu change rather than reformulation with items removed from menus being higher energy than continuous items. Thus, the impact of a calorie labelling policy on food may differ from other policies like the Soft Drinks Industry Levy, which created an economic incentive for, and was associated with substantial, reformulation (31). If food industry actors successfully market out-of-home eating as a treat where customers should indulge in high-calorie options, then business may not be incentivised to reduce the kcal of items offered (14). Evidence from focus groups in the Republic of Ireland found that OOH eating is commonly perceived by families as a treat, with health considered a lower priority (32).

The observation that the reduction in mean kcal was more pronounced when not adjusting for food group (difference of −21 kcal, 95% CI −30 to −12) suggests that the fully adjusted model (difference of −9 kcal, 95% CI −16 to −1) may underestimate the overall effect of the calorie labelling policy. By controlling for food group, our analysis accounts for differences in the types of food items offered before and after the policy implementation. This means that if certain food groups that tend to have lower calorie counts are more represented in the post-policy period, the adjusted model will show a smaller change compared to the model that does not account for these shifts in food group distribution. The policy may have been followed by a shift towards offering more items from lower calorie food groups. For example, our findings showed that new items introduced post-policy had a lower mean calorie count compared to pre-existing items. This suggests that businesses might be strategically altering their menu offerings by including more lower-calorie items, thereby reducing the average calorie content available to consumers. This could be seen as an indirect impact of the calorie labelling policy, where businesses respond by modifying their overall menu composition rather than reformulating existing items.

### Comparison to Previous Literature

Our findings of an overall reduction of 9 kcal pre-post are similar to the magnitude reported in a recent meta-analysis that found mandatory calorie disclosure was associated with a 15 kcal reduction in menu items (11). Similar to our reformulation analysis, more recent work from the USA also found no significant pre-post changes in continuously available items but observed some small reductions in mean kcal due to new items having lower kcal (21). Our study did identify lower mean kcal from new items, but these differences were only significant compared to removed items, not compared to continuous items. Items were similar in total kcal: 445 kcal in the pre-period and 436 kcal in the post-period in our study, and 399 kcal in the pre-period and 388 kcal in the post-period in the USA study (21). Our analysis separates foods into more categories (12 groups instead of 5), allowing for more detailed analysis of where changes occurred. Similar to our findings, a study of menu changes after a mandatory menu labelling legislation in Ontario, Canada did not find evidence of change in mean kcal for continuous or new items (33). We found the largest pre-post changes in pubs, bars and inns; restaurants; and sport and entertainment chains. Previous work from King County, Washington that examined 37 chain restaurants after mandatory calorie labelling was introduced also found greater kcal reductions at full-service restaurants compared to quick-service outlets (34).

### Policy Implications

Overall, we found limited evidence that mandatory calorie labelling in England was associated with significant changes in menu items offered in the out-of-home sector. Alongside our other findings of no pre-post changes in kcal purchased or consumed (12), this suggests that while the policy’s immediate impact may be modest, even small changes can be meaningful at a population level. The limited impact observed may be related to less than perfect implementation, with only 80% of outlets displaying any calorie labelling post-policy and only 15% meeting all implementation requirements (35) along with low intention from local authorities to proactively check implementation in chains (14).

Previous qualitative work identified that large OHFOs within scope of the policy were hesitant to reduce portion size due to concerns around customer satisfaction (14). There may be other strategies that could support customer decisions to select lower calorie options including more actionable contextual information beyond adults’ daily caloric needs. For example, interpretive labels are generally more effective than purely information-based labels in the grocery sector (36). Other potential strategies such as adjusting pricing to make lower calorie options more attractive, employing strategic marketing, or imposing limits on total energy content of individual items may be warranted.

### Strengths and limitations

Our study has several strengths, including the use of the most comprehensive data available on energy content of menu items served by the out-of-home food sector in England that also allows for comparisons of the same chains and food items at both time points. Our efforts to identify removed, new, and continuously available items, as well as examining changes by food type and chain type, allowed us to account for systematic differences in how chains present their items and improve our understanding of where pre-post changes occurred in these data. This large natural experimental analysis examining real-world changes in a diverse set of large OHFOs, may also be generalizable to other country contexts that have similar types of chains.

However, our study also has limitations. Although it is the largest dataset available, MenuTracker includes only menu information from chains that posted kcal information online before and after the policy, thus limiting how representative our findings are of the English out-of-home food environment. Previous work found 256 chains were likely to be subject to the calorie labelling regulations when examined in November 2021 (24). We therefore expect our continuously available sample of 78 chains to cover 30% of the large chain out-of-home food sector that posted nutrition information online at the time of data collection. Given the largest chains with the highest market share are likely to have the most resources to post kcal information online, we would expect our study’s coverage of actual items sold to be larger than the proportion of outlets within scope.

Several limitations relate to the MenuTracker data. Our food group categorizations reflect how items were presented by chains and this may vary from chain to chain. Some chains present mixed dishes as a single item —for example, fish and chips with peas, categorized as a main—whereas others present fish, chips and peas separately, categorised as a main, fried potatoes, and a side. Restaurants were more likely to present mixed dishes as mains, whereas fast food restaurants typically present individual components separately, which explains, in part, why the kcal values were highest at restaurants and pubs, bars, and inns. Furthermore, any systematic changes in how calorie information for mixed dishes was presented by chains pre-post (e.g., reporting kcal information for full meals at pre-policy vs. reporting kcal information for individual components of pre-policy full meals at post-policy) may have impacted our analysis of change over time. We accounted for this issue by adjusting for clustering at the chain type and the individual chain level to account for variations in how chains presented their menus and calculate more accurate estimates of pre-post changes in mean kcal.

Although we assume that posted kcal information is an accurate reflection of the true kcal value of items on menus, the calorie labelling regulations allow for kcal information to be within +/-20% and allow several different methods for estimating energy content (8). In a USA-based restaurant food study, nearly one-fifth of the sample contained over 100kcal more than the stated energy values. However, most energy and nutrient values were consistent with laboratory measurements (37). Any systematic changes in the accuracy of calorie estimates, or method of calculating these, over time may also have impacted our findings.

The uncontrolled before-and-after study design poses a challenge to attributing changes solely to the intervention if there was an ongoing trend in kcal information not related to the policy. However, previous work using MenuTracker data found energy content remained constant from 2018 to 2020 (38). Our analyses are primarily focused on businesses that had kcal information available online pre- and post-policy. However, several businesses did not have kcal information available online prior to the policy, and it is feasible that the policy may have caused some of these businesses to calculate the kcal content of their menu items for the first time and engage in reformulation prior to policy enactment.

### Recommendations for future research

Further research is needed to determine whether there will be greater long-term changes in kcal available at OHFO via gradual reductions. Future research evaluating this policy and future out-of-home food policy evaluations in England may benefit from better surveillance data, longer time series for causal attribution, and linkages to purchase data.

## Conclusion

Our study examined pre-post changes in the energy content of menu items before and after the implementation of the 2022 Calorie Labelling (Out of Home Sector) (England) Regulations, finding a modest reduction of 9 kcal per item. When not adjusting for food group, a larger reduction was observed, suggesting the change is partly influenced by pre-post differences in food group distribution. Changes were primarily driven by the removal of higher-calorie items rather than reformulation of existing items. When analysed by food group, the most significant reductions occurred in beverages, burgers, and mains, indicating that policy impact could be improved if customers select lower kcal items in these categories.

## Data Availability

The codebase is publicly available on GitHub at https://github.com/YuruHuang/MenuTracker.

## Supplementary Materials

**Supplementary Table 1.** Summary statistics (n, %) for each of the 78 core chains categorized by chain type.

| Chain type | n items | % of total |
| --- | --- | --- |
| **Cafes and bakeries** | **7350** | **23.7** |
| Boswell | 684 | 2.2 |
| Benugo Cafe | 187 | 0.6 |
| Boost Juice Bars | 107 | 0.3 |
| Cafe Nero | 502 | 1.6 |
| Coffee #1 | 966 | 3.1 |
| Costa Coffee | 1383 | 4.5 |
| Greggs | 221 | 0.7 |
| Joe & The Juice | 126 | 0.4 |
| PAUL | 91 | 0.3 |
| Pret A Manger | 364 | 1.2 |
| Soho Coffee | 453 | 1.5 |
| Starbucks | 1784 | 5.7 |
| Tesco Cafe | 53 | 0.2 |
| The Cornish Bakery | 25 | 0.1 |
| Thomas the Baker | 45 | 0.1 |
| Tim Hortons | 201 | 0.6 |
| **Western Fast Food & Takeaway** | **7949** | **25.6** |
| Barburrito | 129 | 0.4 |
| Ben & Jerry's | 66 | 0.2 |
| Burger King | 108 | 0.3 |
| Coco Di Mama | 1132 | 3.6 |
| Crussh | 285 | 0.9 |
| Domino's Pizza | 2436 | 7.8 |
| FIVE GUYS | 74 | 0.2 |
| KFC | 298 | 1.0 |
| Krispy Kreme | 50 | 0.2 |
| Leon | 134 | 0.4 |
| McDonalds | 300 | 1.0 |
| Papa John's | 517 | 1.7 |
| Pieminister | 37 | 0.1 |
| Pizza Hut | 598 | 1.9 |
| PizzaExpress | 447 | 1.4 |
| Pure. | 275 | 0.9 |
| Subway | 169 | 0.5 |
| Taco Bell | 328 | 1.1 |
| Tortilla | 27 | 0.1 |
| Tossed | 115 | 0.4 |
| Wimpy | 424 | 1.4 |
| **Pubs Bars Inns** | **7655** | **24.7** |
| All Bar One | 416 | 1.3 |
| Brewhouse and Kitchen | 185 | 0.6 |
| Brewers Fayre | 496 | 1.6 |
| Chef and Brewer | 402 | 1.3 |
| Common Room | 284 | 0.9 |
| Cookhouse & Pub | 431 | 1.4 |
| Ember Inns | 555 | 1.8 |
| Farmhouse Inns | 470 | 1.5 |
| Flaming Grill Pub Co. | 524 | 1.7 |
| Hungry Horse | 555 | 1.8 |
| Marstons | 416 | 1.3 |
| Revolution Vodka Bars | 289 | 0.9 |
| Sizzling Pubs | 745 | 2.4 |
| Tank and Paddle | 172 | 0.6 |
| Town, Pub & Kitchen | 315 | 1.0 |
| Vintage Inns | 497 | 1.6 |
| Walkabout | 177 | 0.6 |
| Wetherspoon | 441 | 1.4 |
| Yate's | 285 | 0.9 |
| **Restaurants** | **5457** | **17.6** |
| Ask | 220 | 0.7 |
| Beefeater Grill | 470 | 1.5 |
| Bella Italia | 394 | 1.3 |
| Cafe Rouge | 304 | 1.0 |
| GBK | 346 | 1.1 |
| Harvester | 813 | 2.6 |
| Loch Fyne | 219 | 0.7 |
| Nandos | 269 | 0.9 |
| Pho | 187 | 0.6 |
| Stonehouse Pizza & Carvery | 592 | 1.9 |
| Table Table | 497 | 1.6 |
| The Real Greek | 71 | 0.2 |
| Toby Carvery | 560 | 1.8 |
| Wagamama | 277 | 0.9 |
| Zizzi | 238 | 0.8 |
| **Sport and Entertainment** | **2065** | **6.7** |
| Cineworld | 524 | 1.7 |
| ODEON | 1134 | 3.7 |
| Vue Entertainment | 407 | 1.3 |
| **Asian Fast Food** | **569** | **1.8** |
| Itsu | 145 | 0.5 |
| Wasabi | 201 | 0.6 |
| YO! Sushi | 223 | 0.7 |
| **Total** | **31045** | **100.0** |

**Supplementary Table 2.** Summary statistics (n, %) for each of the 90 full landscape chains categorized by chain type.

| **Category** | **n items** | **% of total** |
| --- | --- | --- |
| **Cafes and Bakeries** | **7653** | **23.2** |
| AMT Coffee | 38 | 0.1 |
| Asda Cafe | 86 | 0.3 |
| BOSWELL | 684 | 2.1 |
| Benugo Cafe | 187 | 0.6 |
| Boost Juice Bars | 107 | 0.3 |
| Caffe Nero | 502 | 1.5 |
| Coffee #1 | 966 | 2.9 |
| Costa Coffee | 1,383 | 4.2 |
| Greggs | 221 | 0.7 |
| JOE & THE JUICE | 126 | 0.4 |
| Morrisons Cafe | 179 | 0.5 |
| PAUL | 91 | 0.3 |
| Pret A Manger | 364 | 1.1 |
| Sainsbury's Cafe | 158 | 0.5 |
| Soho Coffee | 453 | 1.4 |
| Starbucks | 1,784 | 5.4 |
| Tesco Cafe | 53 | 0.2 |
| The Cornish Bakery | 25 | 0.1 |
| Thomas the Baker | 45 | 0.1 |
| Tim Hortons | 201 | 0.6 |
| **Western Fast Food & Takeaway** | **8043** | **24.4** |
| Barburrito | 129 | 0.4 |
| Ben & Jerry's | 66 | 0.2 |
| Burger King | 108 | 0.3 |
| Chicken Cottage | 66 | 0.2 |
| Coco Di Mama | 1,132 | 3.4 |
| Crussh | 285 | 0.9 |
| Domino's Pizza | 2,436 | 7.4 |
| FIVE GUYS | 74 | 0.2 |
| KFC | 298 | 0.9 |
| Krispy Kreme | 50 | 0.2 |
| Leon | 134 | 0.4 |
| McDonalds UK | 300 | 0.9 |
| Papa John's | 517 | 1.6 |
| Pieminister | 37 | 0.1 |
| Pizza Hut | 598 | 1.8 |
| PizzaExpress | 447 | 1.4 |
| Pure. | 275 | 0.8 |
| Subway | 169 | 0.5 |
| Taco Bell | 328 | 1.0 |
| Tortilla | 27 | 0.1 |
| Tossed | 115 | 0.3 |
| Waterfield's | 28 | 0.1 |
| Wimpy | 424 | 1.3 |
| **Pubs, Bars, Inns** | **8319** | **25.2** |
| All Bar One | 416 | 1.3 |
| Brewers Fayre | 496 | 1.5 |
| Brewhouse and Kitchen | 185 | 0.6 |
| Chef and Brewer | 402 | 1.2 |
| Common Room | 284 | 0.9 |
| Cookhouse & Pub | 431 | 1.3 |
| Ember Inns | 555 | 1.7 |
| Farmhouse Inns | 470 | 1.4 |
| Flaming Grill Pub Co. | 524 | 1.6 |
| Greene King | 20 | 0.1 |
| Hungry Horse | 555 | 1.7 |
| Marstons | 416 | 1.3 |
| Nicholson's | 397 | 1.2 |
| ONeills | 247 | 0.7 |
| Revolution Vodka Bars | 289 | 0.9 |
| Sizzling Pubs | 745 | 2.3 |
| Tank and Paddle | 172 | 0.5 |
| Town, Pub & Kitchen | 315 | 1.0 |
| Vintage Inns | 497 | 1.5 |
| Walkabout | 177 | 0.5 |
| Wetherspoon | 441 | 1.3 |
| Yate's | 285 | 0.9 |
| **Restaurants** | **6242** | **18.9** |
| Ask | 220 | 0.7 |
| Beefeater Grill | 470 | 1.4 |
| Bella Italia | 394 | 1.2 |
| Bills | 169 | 0.5 |
| Browns | 577 | 1.7 |
| Cafe Rouge | 304 | 0.9 |
| GBK | 346 | 1.0 |
| Harvester | 813 | 2.5 |
| Honest Burgers | 39 | 0.1 |
| Loch Fyne | 219 | 0.7 |
| Nandos | 269 | 0.8 |
| Pho | 187 | 0.6 |
| Stonehouse Pizza &<br>Carvery | 592 | 1.8 |
| Table Table | 497 | 1.5 |
| The Real Greek | 71 | 0.2 |
| Toby Carvery | 560 | 1.7 |
| Wagamama | 277 | 0.8 |
| Zizzi | 238 | 0.7 |
| **Sport & Entertainment** | **2155** | **6.5** |
| Cineworld | 524 | 1.6 |
| ODEON | 1,134 | 3.4 |
| Top Golf | 90 | 0.3 |
| VUE ENTERTAINMENT | 407 | 1.2 |
| **Asian Fast Food** | **569** | **1.7** |
| Itsu | 145 | 0.4 |
| Wasabi | 201 | 0.6 |
| YO! Sushi | 223 | 0.7 |
| **Total** | **32,981** | **100.0** |

**Supplementary Table 3.**
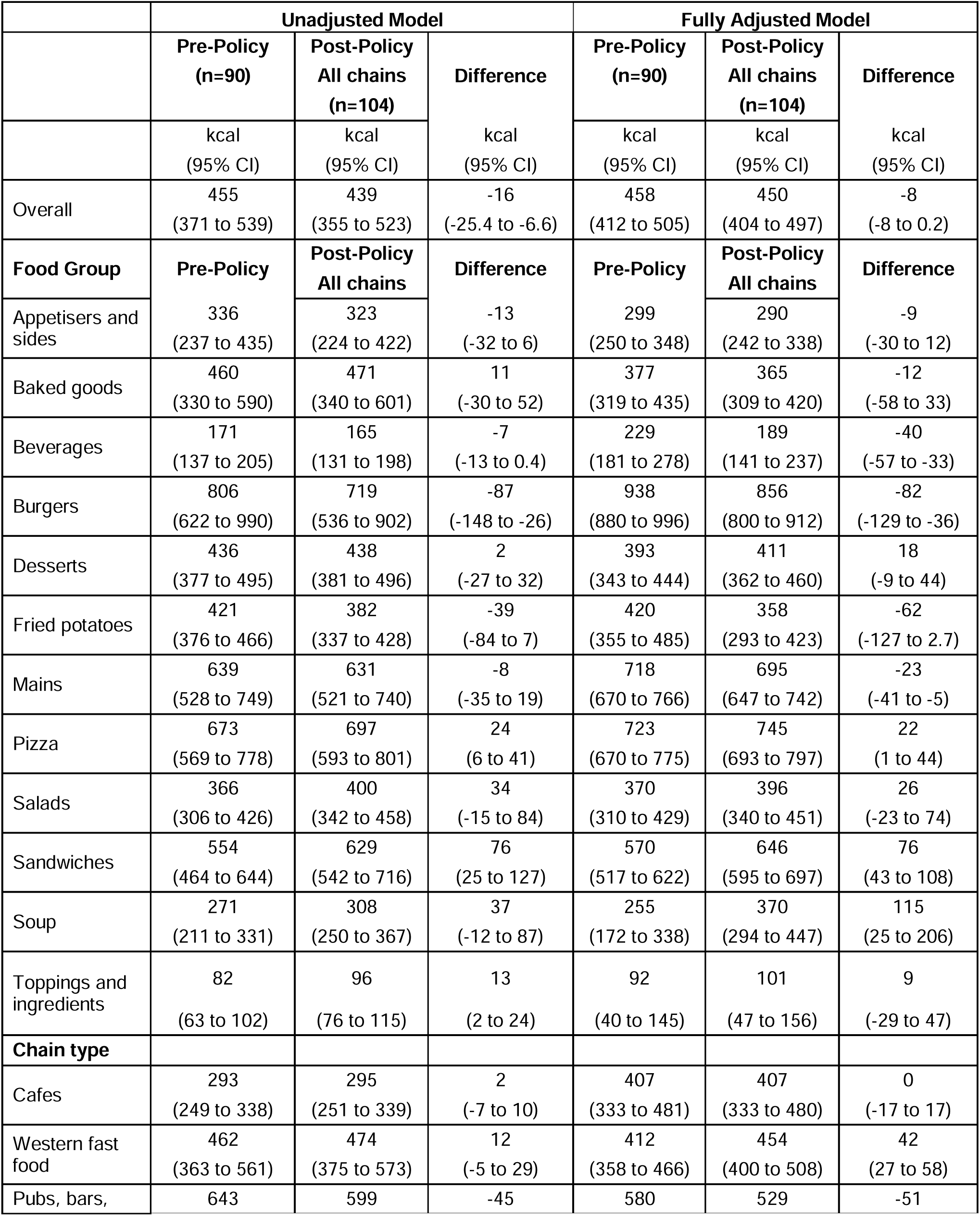
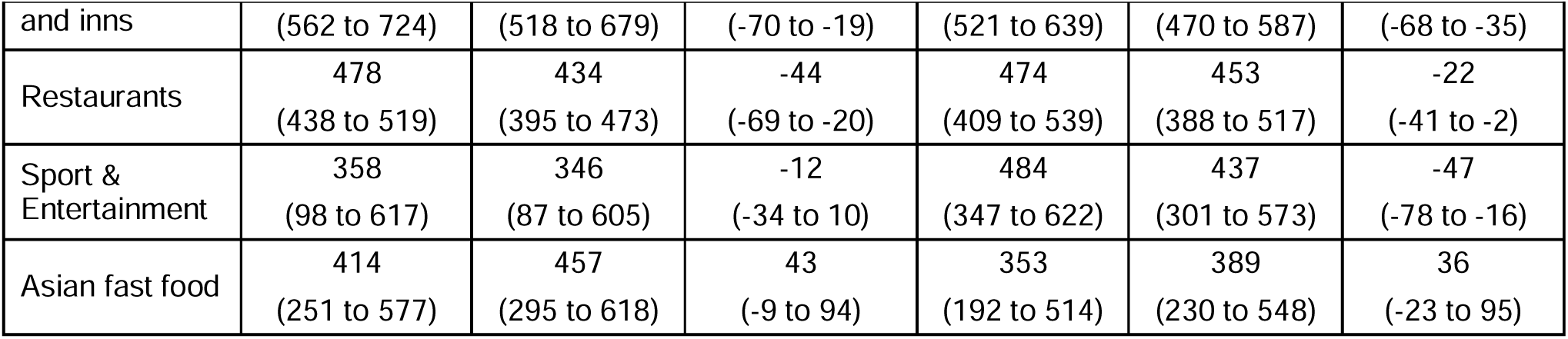
Mean kcal content of all items from all available MenuTracker chains, by food group and by food business type.

## Notes

### Competing Interest Statement

The authors have declared no competing interest.

### Funding Statement

This study is funded by NIHR: Implementation and assessment of mandatory kcal labelling in the out-of-home sector, NIHR200689. The views expressed are those of the authors and not necessarily those of the NIHR or the Department of Health and Social Care.

